# Comparing Methods to Identify which Adult Emergency Department Visits Might be Avoided: A Retrospective Analysis of the MIMIC-IV-ED Database

**DOI:** 10.1101/2025.08.14.25333685

**Authors:** James G. Wrightson, Linda K. Truong, Cobie Starcevich, Piers Truter, Karim M. Khan, Jeffrey Morgan, Kimberlyn McGrail, Clare L. Ardern

**Affiliations:** Department of Family Practice, The University of British Columbia, Vancouver, Canada; Department of Physical Therapy, The University of British Columbia, Surrey, Canada; Centre for Aging SMART, The University of British Columbia, Vancouver, Canada; Curtin School of Allied Health, Curtin University, Perth, Australia; School of Health Sciences, The University of Notre Dame Australia, Perth, Australia; Department of Physiotherapy, Fiona Stanley Hospital, Murdoch, Australia; School of Kinesiology, The University of British Columbia, Vancouver, Canada; School of Population and Public Health, The University of British Columbia, Vancouver, Canada; School of Medicine, Simon Fraser University, Surrey, Canada; Sport & Exercise Medicine Research Centre, La Trobe University, Melbourne, Australia

**Keywords:** “Emergency medicine”, “Triage”, “Healthcare system”

## Abstract

**Introduction:** There are multiple ways to define and identify potentially avoidable emergency department (ED) visits, making it difficult to estimate their frequency accurately. In this study, we compared the proportions and characteristics of potentially avoidable ED visits identified using three commonly used algorithms.

**Methods:** We analysed the publicly available Medical Information Mart for Intensive Care IV Emergency Department dataset to estimate the proportions and characteristics of potentially avoidable ED visits identified using i) the eventual discharge diagnosis (*Diagnosis*), ii) the use of hospital resources (*Resources*), and iii) the triage acuity score assigned to the patient during emergency department triage (*Triage Acuity*).

**Results:** The proportion of visits identified as potentially avoidable differed significantly between algorithms (7 - 36%), and few visits (<2%) were identified as potentially avoidable by all three algorithms. Presenting complaints, discharge diagnoses, and patient demographics were all similarly affected. We found that the proportions and characteristics of potentially avoidable ED visits were affected by the algorithm used to identify them.

**Conclusion:** Methods for identifying potentially avoidable ED visits are not interchangeable. Choosing the appropriate definition and classification method will require researchers to carefully consider the types of visits and the characteristics of patients they wish to identify.

## Introduction

Emergency Departments (EDs) are in crisis. Overcrowding challenges emergency staff who are trying to deliver effective and timely care in the ED [1,2], and detrimentally impacts patient care [3]. It has been widely reported that many ED visits are by patients who do not require treatment in the ED [4–6].

Methods to redirect these patients to appropriate care pathways have been developed, including from the ED to primary care centres [7] or specialists embedded in the ED [8]. Designing and successfully implementing strategies to redirect patients requires accurate population-level estimates of how many and which patients can be redirected. Because there are multiple ways to identify ED visits that might be appropriate for redirection, it is difficult to accurately estimate their frequency [6,9,10]. Inaccurate estimates may lead to misallocation of resources, inadequate primary care capacity, and poorly targeted policy interventions, undermining efforts to reduce ED overcrowding [11]. To identify the resources needed for effective delivery of redirection services, health researchers, health care decision makers, and providers must understand the implications of the algorithm used to identify potentially avoidable ED visits.

ED visits by patients who may not require ED care have been variously described as “non-urgent” [12], “preventable” [13], “non-emergent” [14], “avoidable” [15], “inappropriate” [16], and “low-acuity” [9]. These terms are often used interchangeably, yet they reflect overlapping but distinct paradigms for classifying ED use, may capture different types of ED visits and patients, and are often identified using different algorithms [15]. In this paper, we adopt the terminology proposed by Parkinson et al [15] and describe these visits as “*potentially avoidable ED visits*”. “Avoidable ED visits” is an umbrella term which describes ED visits retrospectively identified using algorithms that can select (i) visits that are divertible to specialised or primary care providers, (ii) visits for preventable conditions (so-called ambulatory care-sensitive conditions), or (iii) visits by patients who did not require medical intervention. As Parkinson et al. suggest, we use the prefix “*potentially”* to emphasise that these algorithms can only identify visits that could, not would, have been avoided [15], and may not reflect the reasons why patients are motivated or required to attend the ED [17,18].

There are multiple methods to classify potentially avoidable ED visits, predominantly using three different types of algorithm: (i) diagnosis-based algorithms, which classify visits/patients according to the patient’s final diagnosis [19–21], (ii) resource use algorithms, which classify visits based on the hospital resources used, including patient admission status [13,22,23] and (iii) triage-based algorithms, which use the urgency or severity assigned at the initial presentation, often quantified by a triage acuity score [24–26]. The language used in the classification methods does not always align intuitively with the operational definition used for potentially avoidable ED visits. For example, researchers have identified “low-acuity” visits using a diagnosis-based algorithm [27] and “non-urgent visits” using an acuity-score-based algorithm [28].

Estimates for the number of potentially avoidable ED visits by adults have been made using the three different classification algorithms. However, these estimates have not been directly compared against each other [29,30]. It is unclear whether the different definitions and classification methods used to identify ED visits affect the frequency, characteristics of visits, and patients identified as potentially avoidable. The aim of this study was to examine the effect of different classification algorithms on characterising potentially avoidable ED visits. We examined how different methods to identify ED visits could affect estimates of how many and which patients and visits are appropriate to redirect from the ED.

## Methods

This study was a cross-sectional retrospective analysis of a publicly available administrative electronic health record dataset. The study was carried out in accordance with the Declaration of Helsinki and received institutional ethics approval from The University of British Columbia (H25-01308). Reporting followed the REporting of studies Conducted using Observational Routinely-collected health Data (RECORD) guidelines [31].

### Data Sources and Participants

The analysis was performed using version 2.2 of the Medical Information Mart for Intensive Care IV (MIMIC-IV) and the MIMIC-IV Emergency Department dataset (MIMIC-ED) [32,33]. MIMIC-ED is a dataset of ∼425,000 de-identified admissions to the Beth Israel Deaconess Medical Center ED (Boston, USA) collected between 2011 and 2019. MIMIC-ED contains data from patients who were admitted to the hospital from the ED, patients who were discharged home after treatment in the ED, and patients who left the hospital voluntarily before receiving care in the ED. The Beth Israel Deaconess Medical Center is an academic medical center, serving neighbourhoods in Boston and surrounding municipalities, which see ∼56,000 patients in the ED per year. During the study period (2011–2019), approximately 98% of state residents had health insurance coverage [34]. The *edstays*, *diagnoses*, and *triage* tables from the MIMIC-ED *ed* module were joined using the *subject_id* and *stay_id* variables. We linked data from MIMIC-ED to the *patients* table from the MIMIC-IV *hosp* module using the *subject_id* variable.

We included data from adults (age at least 18 years) in this study. All visits in which age was not recorded were removed from the sample. To ensure that all three algorithms for identifying potentially avoidable ED visits were compared on an identical sample, visits without a recorded primary diagnosis or triage acuity score were removed. All remaining ED visits between 2011–2019 with complete data were included in the analysis. The study unit was the ED visit. Data processing steps can be found in the online materials.

### Algorithms to Identify Potentially Avoidable ED Visits

We compared three common algorithms for identifying potentially avoidable ED visits:

1. A diagnosis-based algorithm (“*Diagnosis*”) commonly referred to as the New York University Emergency Department algorithm [35,36]. A diagnosis-based algorithm assumes that whether a visit was potentially avoidable can be inferred post-hoc from the discharge diagnosis given to the patient. Full details of the *Diagnosis* algorithm can be found elsewhere [37]. Briefly, the algorithm assigns probabilities to the ICD (9 and 10) primary diagnosis codes across four categories: Non-emergent, Emergent-Primary Care, and two Emergent-ED categories. Primary diagnoses were taken from the ICD-9 or ICD-10 diagnoses assigned by trained coders for billing purposes after discharge. We labelled a visit as potentially avoidable if the summed probability for the Non-emergent and Emergent-Primary Care categories was over 0.5 [37]. Probabilities for the diagnosis codes can be found on the NYU Center for Health and Public Service Research website [38] and in the lists published by Johnston et al [39].
2. A resource-based algorithm (“*Resources*”). Resource-based algorithms assume that whether a visit was potentially avoidable can be inferred from the events that occurred during the ED visit and the disposition (discharge location) [23,40]. The *Resources* algorithm used in this study labels visits where patients were not admitted to the hospital, were not kept in the ED under observation, were discharged home, and did not receive medications while in the ED, as potentially avoidable.
3. A triage acuity algorithm (“*Triage Acuity*”). A triage acuity algorithm uses the triage acuity score assigned to the patient on arrival to the ED and assigns a binary category (Potentially avoidable/Not avoidable) to the patient based on whether they were “low” acuity or not. Triage acuity is recorded in MIMIC-ED using the Emergency Severity Index (ESI) acuity scale, where a higher score represents less acute patients/visits. Visits where patients were assigned either a 4 or 5 on the ESI at triage were labelled as potentially avoidable [41].

Each of the three algorithms was applied to every visit in the analysis sample. Visits could be identified as potentially avoidable by more than one algorithm. Visits were assigned a single label describing which algorithm, combination of algorithms (or no algorithm), identified the visit as potentially avoidable.

### Outcomes

We examined the effect of each classification algorithm on the following outcomes: proportion of ED visits (%) identified as potentially avoidable, most common presenting complaint, three most common discharge diagnoses (rank), patient age (years), sex (female/male, labelled as “gender” in MIMIC-ED), self-reported race, self-reported pain score at triage (/10), acuity assigned at triage (ESI, /5), discharge disposition (Home/Admitted/Other), and length of stay in the ED (minutes) of potentially avoidable ED visits.

### Analysis

Analyses were performed using R (version 4.5.0) [42] and Python (version 3.12.2) [43]. Full analysis details, including a list of the packages and libraries used for analysis, can be found in the online materials.

Summary statistics are reported as either mean [95%CI], median [IQR], or proportion [95%CI]. Because patients could contribute more than one visit, 95% confidence intervals were clustered on patient.

Proportions and means were estimated using intercept-only generalised estimating equations (logistic or Gaussian family, working-independence correlation structure) with robust sandwich standard errors. For proportions, Wald intervals were calculated on the log-odds scale and back-transformed to the proportion scale. When a proportion was 0% or 100%, confidence intervals were estimated using a patient-level cluster bootstrap (2,000 replicates; percentile method). We used an UpSet plot [44], Cohen’s Kappa (*k*) and the positive (*p_pos*) and negative *(p_neg*) agreement (%) [45] to identify the intersections and agreements between the potentially avoidable visits identified using each algorithm. Confidence intervals for Cohen’s kappa and positive and negative agreement were estimated using the same patient-level cluster bootstrap procedure. For the free-text within ‘presenting complaint’, we generated sentence embeddings using the BioBERT sentence-transformer , which were then reduced with principal component analysis (80 components) and clustered using k-means clustering. The number of clusters (k = 75) was selected as the first value for k that was stable across 20 random starts (median adjusted Rand index ≥ 0.80 [46] with a silhouette score above 0.25 [47]. Representative n-grams were extracted from each cluster and summarised for interpretability. Common diagnosed conditions were derived from the primary discharge diagnosis recorded for each visit. Each primary ICD-9-CM or ICD-10-CM code was mapped to its corresponding ICD chapter (for example, diseases of the circulatory system). We used the ICD-9 coding convention to label each chapter; details are provided in Supplemental Material Table S8. We reported the three most frequent ICD chapters as percentages for each group of visits identified by each algorithm.

We performed three sensitivity analyses. In the first, we investigated the effects of removing visits with missing triage acuity scores or age on the proportions of visits identified by the algorithms. In the second, we examined the effects of removing frequent ED users [48] on the proportions of visits identified by the algorithms by including only a single visit per patient. In the third, we examined the effect of using a threshold of 5 for the ESI score used in the *Triage Acuity* algorithm. The methods and results of these analyses are described in the supplemental material.

## Results

We analysed data from 417,723 ED visits from 201,106 patients that occurred between 2011 and 2019. Study population selection is shown in Figure 1. The demographics of the analysis sample are shown in Table 1. The characteristics of the excluded visits are shown in Table S1.

**Figure 1.**
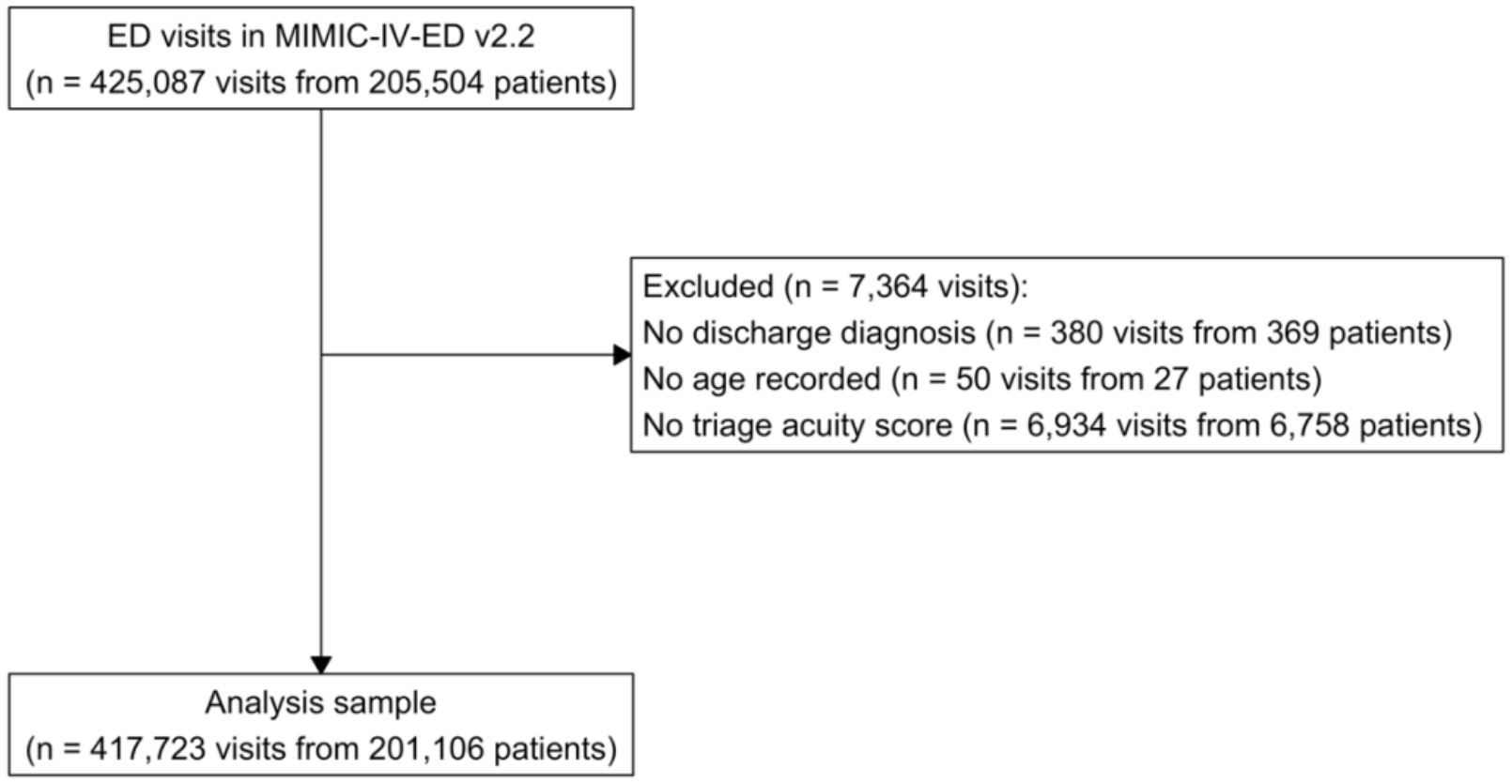
Study population selection

**Table 1.**
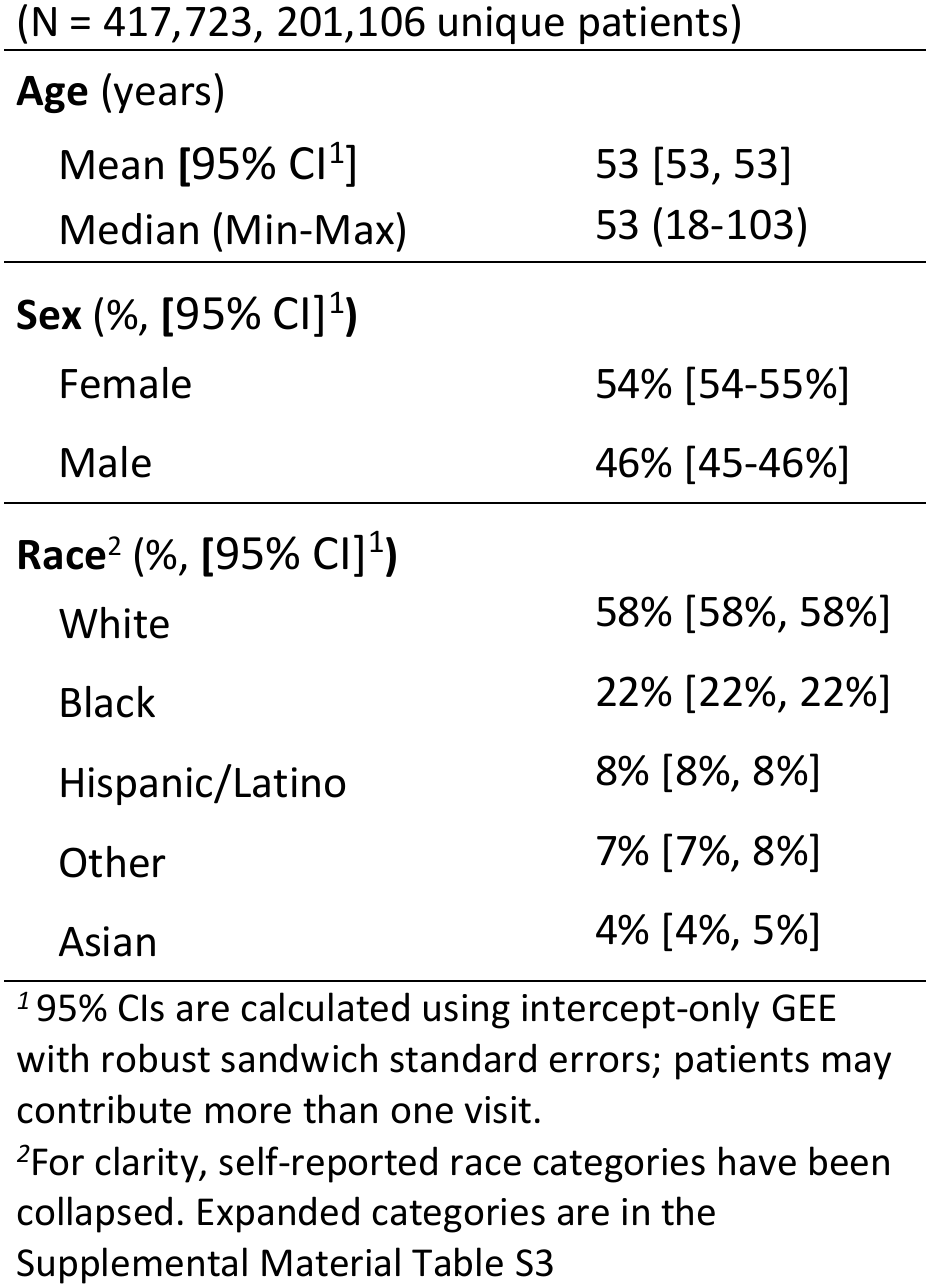
Patient demographics.

| (N = 417,723, 201,106 unique patients) |  |
|---|---|
| **Age** (years) |  |
| Mean **[**95% CI<sup>1</sup>] | 53 [53, 53] |
| Median (Min-Max) | 53 (18-103) |
| **Sex** (%, **[**95% CI]<sup>1</sup>**)** |  |
| Female | 54% [54-55%] |
| Male | 46% [45-46%] |
| **Race**<sup>2</sup> (%, **[**95% CI]<sup>1</sup>**)** |  |
| White | 58% [58%, 58%] |
| Black | 22% [22%, 22%] |
| Hispanic/Latino | 8% [8%, 8%] |
| Other | 7% [7%, 8%] |
| Asian | 4% [4%, 5%] |

<sup>*1*</sup> 95% CIs are calculated using intercept-only GEE with robust sandwich standard errors; patients may contribute more than one visit.
<sup>*2*</sup>For clarity, self-reported race categories have been collapsed. Expanded categories are in the Supplemental Material Table S3

The number of potentially avoidable ED visits that were identified by the algorithms is shown in Figure 2 and Supplemental Material Figure S1. There were 209,429 visits from 121,273 patients identified as potentially avoidable by at least one classification algorithm (≈50% of all ED visits). The characteristics of potentially avoidable ED visits are shown in Table 2 and Supplemental Material Table 2. Few visits (≈2% of all ED visits) were identified as potentially avoidable by all three algorithms. The *Diagnosis* algorithm identified the largest number of unique visits. The Cohen’s Kappa and positive agreement values indicate that agreement was modest between the algorithms (*Diagnosis* x *Resources k* = 0.096 [0.092-0.100], *p_pos* = 34.4 [34.1-34.7], *p_neg* = 73.1 [72.9-73.2], *Diagnosis* x *Triage Acuity k* = 0.048 [0.046-0.050], *p_pos* = 16.1 [15.8-16.3], *p_neg* = 76.9 [76.7-77.0], *Resources* x *Triage Acuity k* = 0.135 [0.131-0.138], *p_pos* = 22.7 [22.4-23.1], *p_neg* = 86.8 [86.7-86.9]).

**Figure 2.**
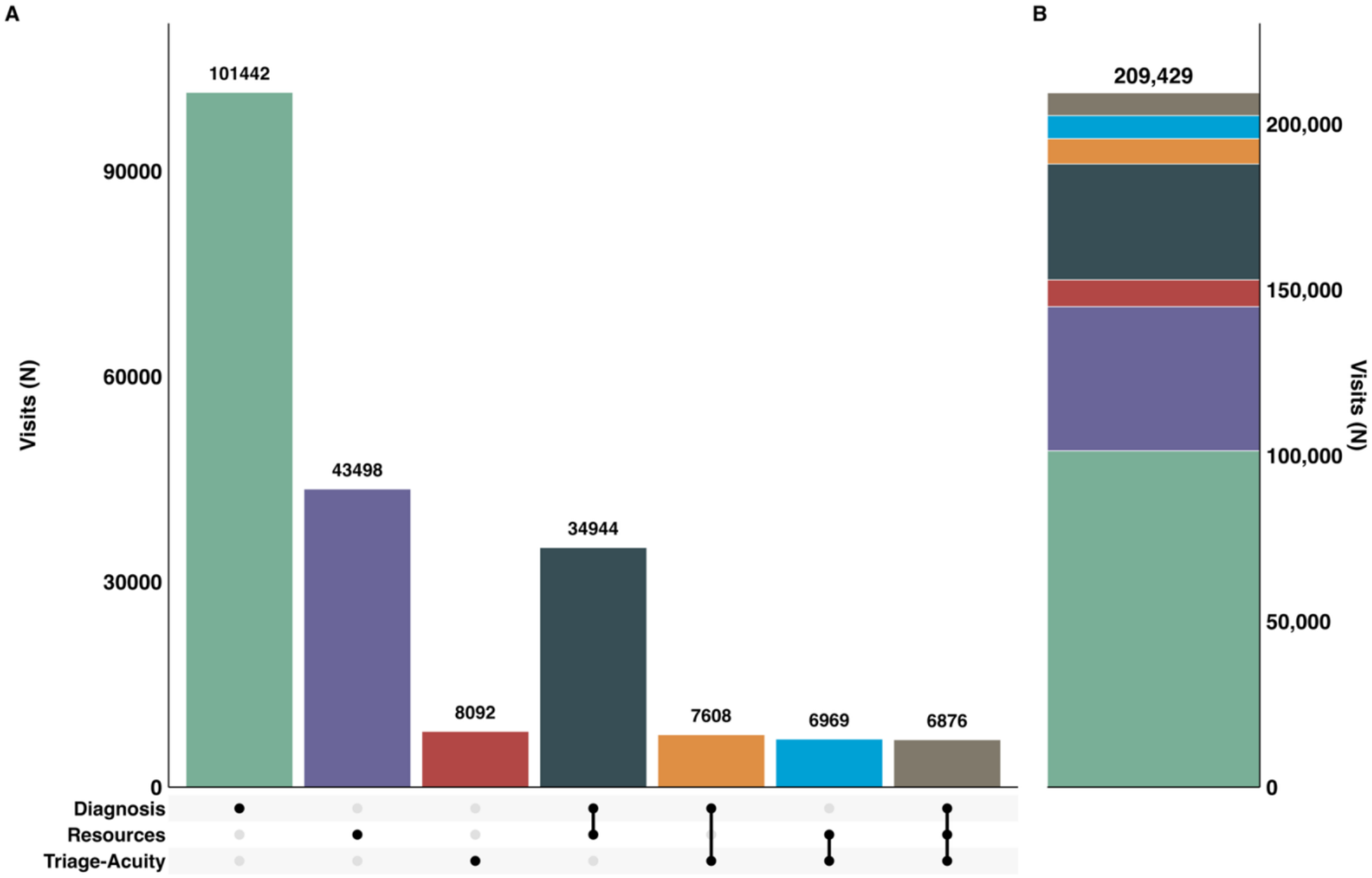
The counts and intersections between the potentially avoidable ED visits identified by each classification algorithm. Panel A (left) shows the intersection of the visits identified with individual and combinations of algorithms. The individual and combinations of algorithms are indicated by single and joined closed circles in the matrix, respectively. The bar displays the number of visits uniquely identified by individual algorithms and combinations of algorithms identified in the matrix. Data are only for visits that were identified by at least one algorithm. Panel B (right) shows the cumulative counts of potentially avoidable visits identified by each algorithm. Data are also shown as a Venn diagram in the supplemental material, Figure S1.

**Table 2.**
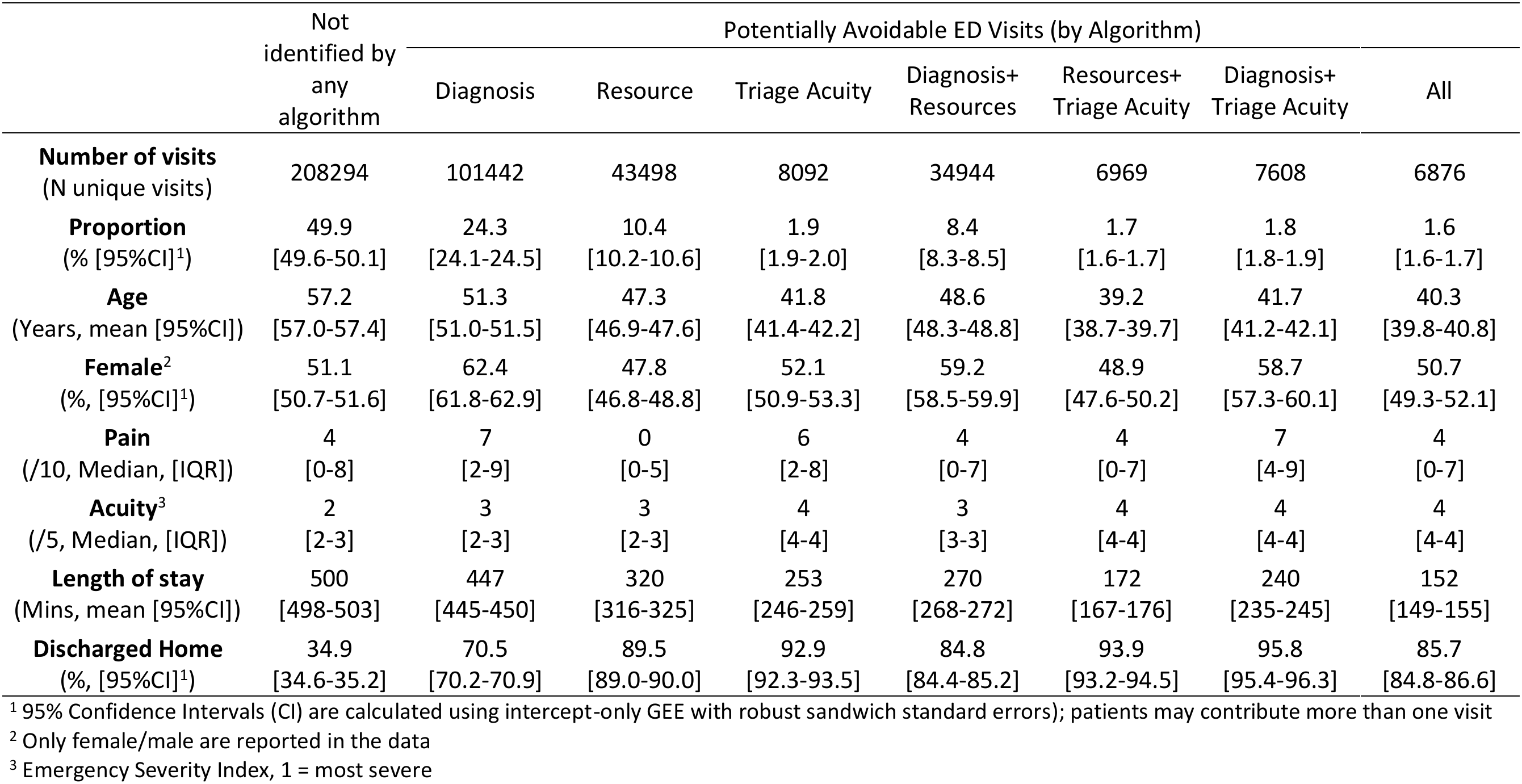
Characteristics of visits not identified as avoidable by any algorithm and visits identified as avoidable by at least one of the algorithms.

Common presenting complaints and conditions represented in the potentially avoidable ED visits are shown in Table 3 and Table 4.

**Table 3.** Common presenting complaints ^1^.

| Rank | Not identified by any algorithm | Potentially Avoidable ED Visits (by algorithm, % of visits) |  |  |  |  |  |  |
|---|---|---|---|---|---|---|---|---|
|  |  | Diagnosis | Resources | Triage Acuity | Diagnosis+<br>Resources | Resources+<br>Triage Acuity | Diagnosis+<br>Triage Acuity | All |
| 1st | Abdomen pain (5.7%) | Nausea vomiting (10.4%) | Alcohol intoxication (11.0%) | Finger laceration (8.9%) | Abdomen pain (8.9%) | Finger laceration (10.4%) | Lower back pain (8.5%) | Rash (9.2%) |
| 2nd | Chest pain (5.3%) | Abdomen pain (10.2%) | Chest pain (5.9%) | Ankle pain (8.1%) | Nausea vomiting (4.9%) | Ankle pain (8.7%) | Back pain (7.5%) | Laceration (6.9%) |
| 3rd | Dyspnea (4.6%) | Chest pain (5.6%) | Syncope (4.7%) | Knee pain (6.6%) | Leg pain (4.5%) | Thumb laceration (6.7%) | Knee pain (6.7%) | Wound evaluation (6.5%) |

<sup>1</sup> From cluster analysis of free-text presenting complaints (see method)

**Table 4.** Common diagnosed conditions^1^.

| Rank | Not identified by any algorithm | Potentially Avoidable ED Visits (by algorithm, % of ED visits) |  |  |  |  |  |  |
|---|---|---|---|---|---|---|---|---|
|  |  | Diagnosis | Resources | Triage Acuity | Diagnosis+<br>Resources | Resources+<br>Triage Acuity | Diagnosis+<br>Triage Acuity | All |
| 1st | Injury and poisoning (22.3%) | Ill-defined conditions (44.5%) | Ill-defined conditions (27.5%) | Injury and poisoning (75.7%) | Ill-defined conditions (52.2%) | Injury and poisoning (72.9%) | Diseases of the MSK system (45.7%) | Diseases of the MSK system (28.5%) |
| 2nd | Diseases of the circulatory system (12.5%) | Diseases of the MSK system (15.5%) | Injury and poisoning (27.3%) | Diseases of the MSK system (5.1%) | Diseases of the MSK system (17.6%) | Mental disorders (5.8%) | Ill-defined conditions (10.8%) | Factors influencing health status/contact with health services (15.3%) |
| 3rd | Diseases of the digestive system (11.4%) | Diseases of the skin and subcutaneous tissue (7.3%) | Mental disorders (20.2%) | Mental disorders (3.2%) | Diseases of the genitourinary system (4.8%) | Factors influencing health status/contact with health services (4.2%) | Diseases of the skin and subcutaneous tissue (10.2%) | Ill-defined conditions (14.6%) |

MSK; Musculoskeletal
<sup>1</sup> From the ICD-9-CM and ICD-10-CM chapters for the primary diagnosis ICD code, see Supplemental Material Table S8

## Discussion

Our results suggest that methods for identifying potentially avoidable ED visits are not interchangeable: the choice of the algorithm determines how many and which patients and conditions are identified as potentially avoidable. There was modest agreement between the different methods used to identify potentially avoidable ED visits, and few visits were identified as potentially avoidable by all three classification algorithms. There were differences between the algorithms for the numbers and proportions of visits, the characteristics of the visits, and patients. Choosing the appropriate definition and classification method will require researchers to carefully consider the types of visits and the characteristics of patients they wish to identify.

Our results extend previous findings in adults [29,30] and infants [49] by showing that classification methods not only influence estimates of the frequency and proportion of potentially avoidable ED visits, but also affect the types of patients, conditions, and visits identified as potentially avoidable. The *Diagnosis* algorithm identified the greatest number of potentially avoidable ED visits. The greatest overlap in identified visits occurred between the *Diagnosis* and *Resources* algorithms, but even this intersection was smaller than the number of visits identified by either algorithm individually. In comparison, few visits were identified solely by the *Triage Acuity* algorithm. Accurate estimates of the number of patients or visits, and the type of care are essential for effective health care planning and health system needs assessments [50]. Although there is increasing interest in developing methods to reduce potentially avoidable ED visits, the strength of the evidence supporting these approaches is unclear, in part because of the heterogeneity in how these visits are defined and identified [1]. Adopting a definition and taxonomy similar to what Parkinson et al. [15] propose could provide different classes of potentially avoidable visits against which different algorithms could be validated. However, at present there is no consensus on how to define potentially avoidable ED visits [6]. Failing to define potentially avoidable ED visits (i.e. what researchers and policy makers want to measure) can result in confusion between the type of ED visit of interest and the algorithm (and thus operational definition) used to identify the visit (e.g. examining the validity of the discharge diagnosis to identify “low-acuity” ED visits) [21]. Valid measurement is only possible when the measurement object is clearly defined [51], and the development of an agreed taxonomy for potentially avoidable visits is a necessary next step for researchers and policy makers in this field.

In our analysis, the median triage acuity score for potentially avoidable visits identified with either the *Diagnosis* or *Resources* algorithms (3/5) was higher than the score (4/5) typically used as a cut-off for the *Triage Acuity* algorithm. Compared with *Diagnosis*- or *Resource-based* methods, there was little benefit in using *Triage Acuity* alone to retrospectively identify potentially avoidable ED visits. However, it is the only algorithm that can be deployed *during* the ED visit, and the only one that reflects how the patient presented to the ED and the clinician’s predictions about the type and urgency of care the patient would require. The *Diagnosis-* and *Resource-based* algorithms can only be used retrospectively—an obvious limitation. Retrospectively comparing the differences between visits triaged as low-acuity with those labelled as potentially avoidable may provide insight into how potentially avoidable visits could be more effectively identified at the time of the ED visit. This is an essential step before one could effectively redirect a patient to more appropriate care away from the ED [7]. The relatively large disparities between the numbers of unique and shared visits identified by the *Diagnosis* and *Resource-*based algorithms indicate that one cannot assume the estimates for the numbers of potentially avoidable ED visits identified with one algorithm represent the numbers of visits identified by the other.

The choice of algorithm, in effect, determines the frequency and types of patients, conditions, and visits that could be considered potentially avoidable. The definitions are not interchangeable, and we suggest that the validity of the definitions is uncertain. This is important because the choice of algorithm has direct implications for health care planning and resource use. For instance, patients identified by the three algorithms presented with different complaints, reported varying pain levels at triage, received distinct diagnoses, and were hospitalised in differing numbers. Researchers, health care administrators, and policy makers should carefully consider the definitions and assumptions underlying each algorithm when selecting one to identify potentially avoidable ED visits.

When attempting to identify the types of services that can be implemented and the resources needed for effective delivery, healthcare providers and health researchers must carefully consider the implications of the algorithm used to identify potentially avoidable ED visits. For example, automated methods to predict which visits are avoidable (e.g., [52,53]) may benefit from examining how their models perform across different algorithms. It may also be necessary to develop new algorithms for specific use cases (e.g., redirecting patients with low back pain from the ED to a specialist clinic within the same hospital). An algorithm developed to identify patients who are suitable for redirection might require combining the eventual diagnoses, expected hospital resource use, and the severity as estimated at triage. Validation of these algorithms should, based on our results, involve comparing the results from new algorithms with those from existing algorithms to understand differences in prevalence and patient characteristics, and subsequent health care use and costs. These types of analyses could inform future efforts to refine existing algorithms and improve how truly avoidable ED visits are identified. Similarly, healthcare providers, administrators or decision makers wishing to estimate how many patients are expected to be redirected to specialist clinics may wish to combine algorithms to ensure they identify not only the correct conditions (a *Diagnosis* algorithm) but also the specific resources required.

It has long been suspected that the criteria used to define and identify potentially avoidable ED visits may not be interchangeable [29,30,49]. One solution to the disparity in definitions and methods used to identify potentially avoidable ED visits is to develop a gold standard case definition [15,40]. This approach assumes there is a true class of ED visits that are potentially avoidable/non-urgent/non-emergent. However, it may be more appropriate to assume that there are different categories of potentially avoidable ED visits, each reflecting different conditions and health care requirements, and that can each be operationally defined in different ways, depending on the goals of the researcher and the characteristics of the data and source of the data [15]. Therefore, an alternative, but less widely adopted, approach would be to adopt a utilitarian perspective and assume that the disparity between algorithms has utility. With this approach, researchers and health planners and administrators would treat each algorithm as a means of classifying different types of potentially avoidable ED visits and patients. For example, if the primary purpose was to measure the frequency of patients with certain conditions who could be redirected from the ED, a diagnosis-based algorithm might be more suitable than if the purpose was to identify patients who would not be admitted. However, if both outcomes are important, then combining algorithms may be preferable to, for example, identifying patients with pre-defined conditions who are less likely to use significant hospital resources. By comparing the results of multiple algorithms, researchers, health planners and administrators, and policymakers can make an informed decision about how to amend or combine algorithms to identify the visits and patients they intend to identify.

A limitation of our study, and more generally of the oft-used definitions for potentially avoidable ED visits, is that they do not reflect the complexity of ED visits and clinical presentations [54]. For example, a 90-year-old patient with a musculoskeletal condition classified as potentially avoidable by the *Diagnosis* algorithm may still require ED care, whereas a 20-year-old with the same condition may not. Similarly, visits by patients who are diagnosed with mental, behavioural, or neurodevelopmental disorders (one of the most common diagnoses identified by the *Resource* algorithm) may not use many ED resources, but attending the ED may be a necessary first step to accessing care. Each algorithm has its own inherent limitations. For the *Diagnosis* algorithm, initial severity (and thus whether the visit was avoidable) may not be reflected in the final diagnosis, and variability in how institutions and individuals assign ICD diagnosis codes [55] may affect the generalizability of the findings for that algorithm. The *Resource* and *Triage* algorithms are affected by local practice patterns and staff training, which may be exacerbated by the single center data source. We encourage researchers and health planners and administrators to consider a priori which conditions and patients should *not* be classified as potentially avoidable, and refine their choice of algorithm.

Because the data we analysed are from a single center, the types of patients (e.g. there are no data from paediatric patients in the dataset), the number of visits, severity/acuity distribution and missingness at this institution may not reflect those encountered in other ED settings. For example, ESI 4 and 5 visits (i.e. potentially avoidable) account for approximately 20–30% of visits captured in the National Hospital Ambulatory Medical Care Survey (NHAMCS) [40], compared to a lower proportion in our sample, consistent with the study center’s role as an academic tertiary referral center in a well-resourced primary care market. Therefore, our findings may be susceptible to spectrum bias. Replication of these findings in other datasets, particularly from other countries and health systems, is warranted. It is not clear whether the characteristics of patients and visits reported here would generalise to other health systems (especially those that use other triage acuity scales) and to paediatric patients. However, we expect the disparity between algorithms to be evident in similar health systems (e.g., [29,30,49]).

## Conclusions

The choice of algorithm influenced the number and types of visits and patients identified as potentially avoidable ED visits. Rather than using a single algorithm to identify potentially avoidable ED visits, we encourage researchers and health systems to develop or refine algorithms that reflect the purpose and priorities of their own analyses. We encourage analysts to adopt a utilitarian view of the disparities between different algorithms by contrasting results derived from different algorithms.

## Data Availability

MIMIC-ED is publicly available from Physionet. Access to MIMIC-ED is granted only after completing the Collaborative Institutional Training Initiative. California health data is publicly available from the California Health and Human Services Open Data Portal. The codebooks for this study are available online

https://physionet.org/

https://data.chhs.ca.gov/

https://github.com/James-G-Wrightson/mimic_msk

## List of Abbreviations

BioBERT: Bidirectional Encoder Representations from Transformers for Biomedical Text Mining
ED: Emergency Department
ESI: Emergency Severity Index
ICD: International Classification of Diseases
MIMIC-ED: Medical Information Mart for Intensive Care IV Emergency Department dataset

## Declarations

### Ethics Approval and Consent to Participate

This research was conducted in accordance with the Declaration of Helsinki and received institutional ethics approval from The University of British Columbia (H25-01308). This study uses deidentified data from the publicly available MIMIC-IV database. The original curation of MIMIC-IV was approved by the Institutional Review Board at Beth Israel Deaconess Medical Center which granted a waiver of informed consent, and thus no new consent from participants was required for this study.

### Availability of Data and Materials

The MIMIC-ED data are publicly available from Physionet [48]. Code is available online: https://github.com/James-G-Wrightson/mimiced_compare.git

### Competing Interests

The authors declare that they have no competing interests.

### Funding

None

### Authors’ Contributions

JGW: Conceptualisation, Methodology, Investigation, Writing – Original Draft Preparation. LKT: Writing – Review & Editing. CS: Writing – Review & Editing. PT: Writing – Review & Editing. KMK: Writing – Review & Editing. JM: Writing – Review & Editing. KM: Writing – Review & Editing. CLA: Conceptualisation, Methodology, Writing – Review & Editing.

## Acknowledgements

Not applicable

## AI Disclosure Statement

The authors used Claude Fable 5 and Claude Code (Anthropic, PBC) to edit code used for analysis and producing figures in response to reviewer comments during peer review. The output was reviewed and verified by the authors.

## Notes

### Competing Interest Statement

The authors have declared no competing interest.

### Author Declarations

The study received institutional ethics approval from the University of British Columbia Behavioural Research Ethics Board (H25-01308).

### Summary of Updates

These revisions respond to reviewer feedback and are significant enough to warrant a new version of the preprint. The major change is that the unit of study is now the visit, rather than the patient. By-patient analyses are moved to the Supplemental Material. Other changes: Study flow diagram added (Figure 1). Confidence intervals account for repeat visits. Positive and negative agreement now reported alongside Cohens kappa. Presenting-complaint clustering redone (k = 75). Table 3 was updated to match. Results tables restructured. Tables 2 to 4 cover every combination of algorithms and use visits not identified by any algorithm as the comparator. Methods clarified: the source of diagnosis codes, how visits flagged by more than one algorithm are labelled, and the sources of the diagnosis-code probabilities.Three sensitivity analyses (Supplemental Material): missing-data exclusions, one visit per patient, and Triage-Acuity defined as ESI 5 only. Supplemental Material additions: an overlap Venn diagram (Figure S1), dispositions (Table S2), expanded race categories (Table S3), and the ICD chapter mapping (Table S8).

