## Supplemental Material for "Comparing Methods to Identify which Adult Emergency Department Visits Might be Avoided: A Retrospective Analysis of the MIMIC-IV-ED Database"

### Figure S1

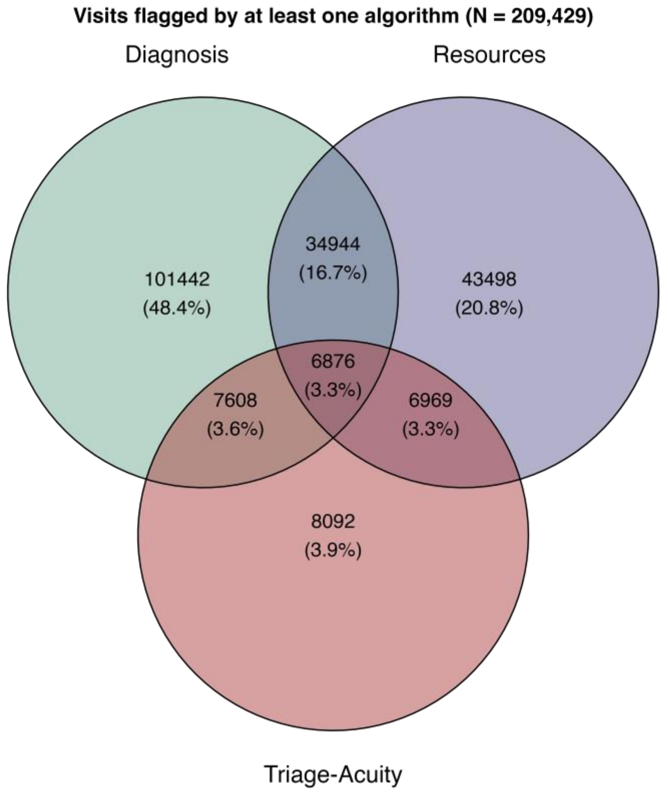


**Figure S1.** Overlap of ED visits identified as potentially avoidable by the Diagnosis, Resources, and Triage-Acuity algorithms. Visits flagged by at least one algorithm (N = 209,429; 50.1% of the analysis sample, N = 417,723). Percentages are of flagged visits.

### Table S1

| **Table S1**. ICD-9 and ICD-10 Chapters and labels for the common presenting complaint | | |
|---|---|---|
| Chapter | ICD-9 | ICD-10 |
| Infectious and parasitic diseases | 001-139 | A00-B99 |
| Neoplasms | 140-239 | C00-D49 |
| Diseases of the blood and blood-forming organs | 280-289 | D50-D89 |
| Endocrine, nutritional and metabolic diseases | 240-279 | E00-E99 |
| Mental disorders | 290-319 | F00-F99 |
| Diseases of the nervous system | 320-389 | G00-G99 |
| Diseases of the circulatory system | 390-459 | I00-I99 |
| Diseases of the respiratory system | 460-519 | J00-J99 |
| Diseases of the digestive system | 520-579 | K00-K99 |
| Diseases of the skin and subcutaneous tissue | 680-709 | L00-L99 |
| Diseases of the musculoskeletal system | 710-739 | M00-M99 |
| Diseases of the genitourinary system | 580-629 | N00-N99 |
| Pregnancy, childbirth and the puerperium | 630-679 | O00-O99 |
| Certain conditions originating in the perinatal period | 760-779 | P00-P99 |
| Congenital anomalies | 740-759 | Q00-Q99 |
| Ill-defined conditions | 780-799 | R00-R99 |
| Injury and poisoning | 800-999 | S00-T99 |
| Factors influencing health status/contact with health services | V codes | Z00-Z99 |
| Other | All remaining codes | All remaining codes |

### Table S2

Table S2. Proportions of visits with each disposition identified by the different algorithms

| Disposition | Not identified by any algorithm | Diagnosis | Resources | Triage-Acuity | Diagnosis + Resources | Resources + Triage-Acuity | Diagnosis + Triage-Acuity | All |
|---|---|---|---|---|---|---|---|---|
| Home | 208,294 | 101,442 | 43,498 | 8,092 | 34,944 | 6,969 | 7,608 | 6,876 |
| Admitted | 72,716 | 71,562 | 38,925 | 7,520 | 29,637 | 6,542 | 7,291 | 5,894 |
| Transfer | 125,937 | 27,026 | NA[1] | 409 | NA[1] | NA[1] | 158 | NA[1] |
| Left without being seen | 5,705 | 1,152 | NA[1] | 69 | NA[1] | NA[1] | 36 | NA[1] |
| Eloped | 47 | 74 | 1,525 | 3 | 3,519 | 209 | 10 | 677 |
| Left against medical advice | 1,151 | 884 | 1,878 | 70 | 1,179 | 175 | 92 | 218 |
| Expired | 645 | 459 | 437 | 7 | 291 | 4 | 13 | 8 |
| Other | 170 | 24 | NA[1] | 1 | NA[1] | NA[1] | 2 | NA[1] |

[1]NA; Not applicable. The Resources algorithm excludes visits that were admitted, transferred, or that died in the ED, by design

95% CIs are calculated using intercept-only GEE with robust sandwich standard errors; 0% cells use a patient-level bootstrap.

### Table S3

Table S3. Mapping of self-reported race categories in MIMIC-IV-ED to the collapsed groups used in Table 1

| MIMIC-IV-ED race category | Collapsed group | Visits (n) | Visits (%) |
|---|---|---|---|
| WHITE | White | 224,797 | 53.8 |
| WHITE - OTHER EUROPEAN | White | 8,912 | 2.1 |
| WHITE - RUSSIAN | White | 6,016 | 1.4 |
| WHITE - BRAZILIAN | White | 1,464 | 0.4 |
| WHITE - EASTERN EUROPEAN | White | 1,297 | 0.3 |
| BLACK/AFRICAN AMERICAN | Black | 76,028 | 18.2 |
| BLACK/CAPE VERDEAN | Black | 7,585 | 1.8 |
| BLACK/AFRICAN | Black | 4,833 | 1.2 |
| BLACK/CARIBBEAN ISLAND | Black | 3,619 | 0.9 |
| HISPANIC/LATINO - PUERTO RICAN | Hispanic/Latino | 13,897 | 3.3 |
| HISPANIC/LATINO - DOMINICAN | Hispanic/Latino | 8,254 | 2.0 |
| HISPANIC OR LATINO | Hispanic/Latino | 3,066 | 0.7 |
| HISPANIC/LATINO - GUATEMALAN | Hispanic/Latino | 2,316 | 0.6 |
| HISPANIC/LATINO - SALVADORAN | Hispanic/Latino | 1,480 | 0.4 |
| HISPANIC/LATINO - COLUMBIAN | Hispanic/Latino | 1,296 | 0.3 |
| HISPANIC/LATINO - MEXICAN | Hispanic/Latino | 1,251 | 0.3 |
| HISPANIC/LATINO - HONDURAN | Hispanic/Latino | 995 | 0.2 |
| HISPANIC/LATINO - CENTRAL AMERICAN | Hispanic/Latino | 780 | 0.2 |
| HISPANIC/LATINO - CUBAN | Hispanic/Latino | 778 | 0.2 |
| ASIAN - CHINESE | Asian | 7,265 | 1.7 |
| ASIAN | Asian | 7,209 | 1.7 |
| ASIAN - ASIAN INDIAN | Asian | 1,547 | 0.4 |
| ASIAN - SOUTH EAST ASIAN | Asian | 1,510 | 0.4 |
| ASIAN - KOREAN | Asian | 773 | 0.2 |
| OTHER | Other | 20,334 | 4.9 |
| UNKNOWN | Other | 5,427 | 1.3 |
| PORTUGUESE | Other | 1,426 | 0.3 |
| SOUTH AMERICAN | Other | 1,054 | 0.3 |
| AMERICAN INDIAN/ALASKA NATIVE | Other | 1,019 | 0.2 |
| PATIENT DECLINED TO ANSWER | Other | 599 | 0.1 |
| NATIVE HAWAIIAN OR OTHER PACIFIC ISLANDER | Other | 488 | 0.1 |
| MULTIPLE RACE/ETHNICITY | Other | 272 | 0.1 |
| UNABLE TO OBTAIN | Other | 136 | 0.0 |

A category is assigned to White, Black, Hispanic/Latino or Asian if its MIMIC-IV-ED label contains that keyword (WHITE, BLACK, HISPANIC or ASIAN); every other label, including Unknown, Patient declined to answer, Unable to obtain, and multiracial or other labels, is assigned to Other. Percentages are of all visits in the analysis sample.

### Sensitivity analysis 1 — visits excluded for missing data

This sensitivity analysis explored the effect of removing the ED visits with missing age, primary diagnosis or triage acuity score data.

**Table S4.** Characteristics of the visits included in the analysis sample and the excluded visits

| **Characteristic** | **Analysis sample** N = 417,723 | **Excluded visits** N = 7,364 | **Difference (95% CI)** [1] |
|---|---|---|---|
| **Age (years)** | | | |
| Mean (SD) | 52.8 (20.6) | 54.2 (21.4) | -1.3 [-1.8, -0.8] |
| **Gender** | | | |
| Female | 226,798 (54%) | 3,100 (42%) | +12.2 [11.0, 13.4] |
| Male | 190,925 (46%) | 4,264 (58%) | -12.2 [-13.4, -11.0] |
| **Self-reported race** | | | |
| White | 242,486 (58%) | 3,516 (48%) | +10.3 [9.1, 11.5] |
| Black | 92,065 (22%) | 933 (13%) | +9.4 [8.5, 10.2] |
| Hispanic/Latino | 34,113 (8.2%) | 398 (5.4%) | +2.8 [2.2, 3.3] |
| Asian | 18,304 (4.4%) | 224 (3.0%) | +1.3 [0.9, 1.7] |
| Other | 30,755 (7.4%) | 2,293 (31%) | -23.8 [-24.9, -22.7] |
| **Pain score (0-10)** | | | |
| Mean (SD) | 4.3 (3.8) | 4.2 (4.0) | +0.1 [-0.3, 0.5] |
| **Length of stay (minutes)** | | | |
| Mean (SD) | 429 (370) | 272 (263) | +157 [151, 163] |
| **Discharge disposition** | | | |
| Admitted | 153,530 (37%) | 4,480 (61%) | -24.1 [-25.2, -22.9] |
| Died in the ED | 197 (<0.1%) | 180 (2.4%) | -2.4 [-2.7, -2.0] |
| Home | 240,087 (57%) | 1,545 (21%) | +36.5 [35.5, 37.5] |
| Other | 23,909 (5.7%) | 1,159 (16%) | -10.0 [-10.9, -9.2] |

[1] 95% confidence interval (CI) calculated using intercept-only GEE with robust sandwich standard errors

There were 201,106 unique patients in the analysis sample, and 7,134 unique patients in the excluded visits. Excluded visits differed systematically: they had a higher proportion of male patients and a higher proportion of patients who self-reported their race as “Other” (31% vs 7.4%). A greater proportion of excluded visits were admitted or died in the ED.

| **Table S5.** Potentially avoidable visit categories: analysis sample compared with every raw ED visit. | | | |
|---|---|---|---|
| Avoidable visit category | Analysis sample N = 417,723, % (n) | All ED visits[1] N = 425,087, % (n) | Difference |
| Diagnosis only | 24.3 (101,442) | 23.9 (101,767) | +0.3 |
| Resources only | 10.4 (43,498) | 10.4 (44,321) | -0.0 |
| Triage-Acuity only | 1.9 (8,092) | 1.9 (8,114) | +0.0 |
| Diagnosis + Resources | 8.4 (34,944) | 8.3 (35,350) | +0.0 |
| Resources + Triage-Acuity | 1.7 (6,969) | 1.6 (7,003) | +0.0 |
| Diagnosis + Triage-Acuity | 1.8 (7,608) | 1.8 (7,608) | +0.0 |
| All three | 1.6 (6,876) | 1.6 (6,879) | +0.0 |
| None | 49.9 (208,294) | 50.4 (214,045) | -0.5 |

[1] All ED visits = Analysis sample+ Excluded visits

Including the excluded visits in the analysis made little difference in the proportions or counts of visits inedited as potentially avoidable by at least one algorithm.

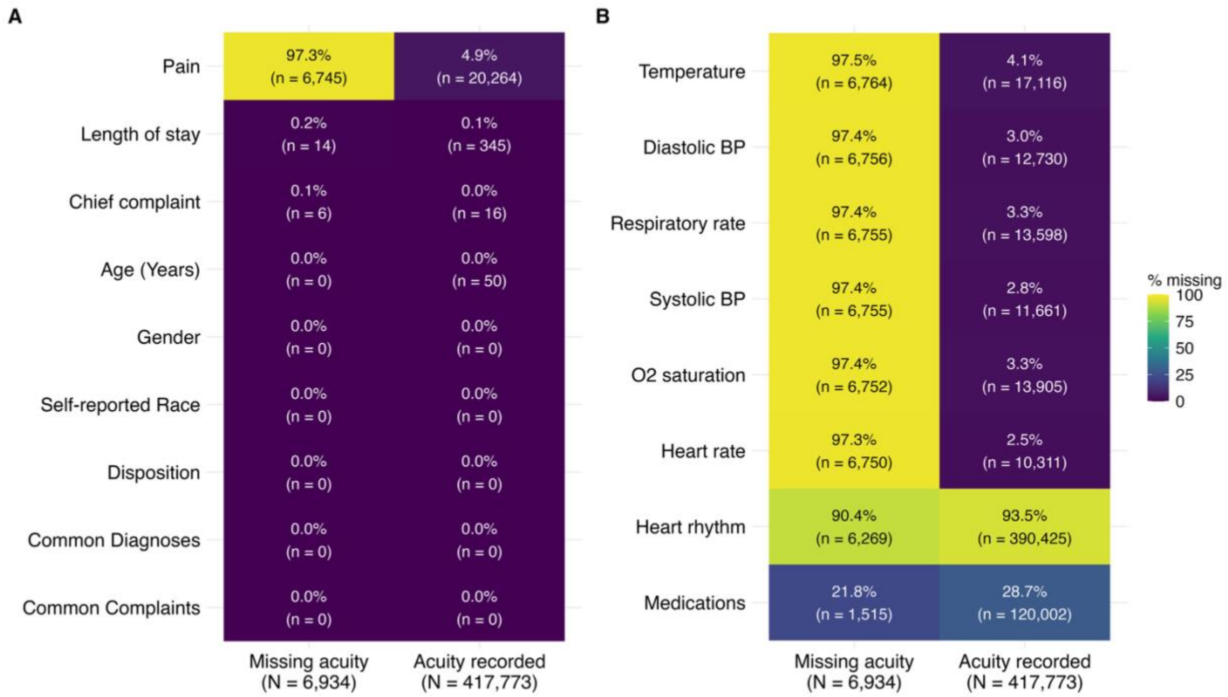


Figure S2. Missingness for variables analyzed in the study (A) and for measurements taking during triage but not analyzed in this study (B)

Missingness patterns suggest that missing triage acuity scores were a consequence of factors that occurred during triage, because they co-occur with missingness in other variables recorded at triage such as pain and heart rate

### Sensitivity analysis 2 — By patient analysis

In the main analysis, the unit of study was the visit. Patients could contribute more than one visit, and there were 216,617 repeat visits in the analysis sample. To examine whether frequent ED users may have biased the results, this sensitivity analysis examined the effect of reducing the analysis sample to include only one visit per patient by including only the first visit of each patient in the analysis sample.

| **Table S6.** Characteristic of the sensitivity analysis sample (N = patients/visits)[1] | |
|---|---|
| **Age (Years, mean)** | 49 [49, 49] |
| (Years, median) | 49 (18-102) |
| **Sex (%)** | |
| Female | 54% [53%, 54%] |
| Male | 46% [46%, 47%] |
| **Self-reported Race (%)[2]** | |
| White | 62% [62%, 62%] |
| Black | 16% [16%, 16%] |
| Other | 10% [9.7%, 10%] |
| Hispanic/Latino | 6% [6.4%, 6.6%] |
| Asian | 6% [5.5%, 5.7%] |

[1]Confidence interval (CI) for categorical variables is computed using Agresti-Coull interval.
[2]For clarity, self-reported races have been collapsed.
Abbreviation: CI = Confidence Interval

**Table S7.** Potentially avoidable visit categories in the one-visit-per-patient cohort

| | Not identified by any algorithm | Potentially Avoidable ED Visits (by Algorithm) | | | | | | |
|---|---|---|---|---|---|---|---|---|
| | | Diagnosis | Resource | Triage Acuity | Diagnosis+ Resources | Resources+ Triage Acuity | Diagnosis+ Triage Acuity | ALL |
| Number of visits (N) | 100,538 | 44,119 | 23,427 | 4,690 | 17,306 | 4,278 | 3,578 | 3,170 |
| Proportion (%, [95% CI]) | 50.0 [49.8-50.2] | 21.9 [21.8-22.1] | 11.6 [11.5-11.8] | 2.3 [2.3-2.4] | 8.6 [8.5-8.7] | 2.1 [2.1-2.2] | 1.8 [1.7-1.8] | 1.6 [1.5-1.6] |
| Age (years, mean [95% CI]) | 54.4 [ 54.3-54.6] | 47.0 [46.9-47.2] | 42.2 [42.0-42.5] | 38.1 [37.6-38.5] | 44.5 [44.2-44.8] | 35.3 [34.9-35.8] | 37.8 [37.3-38.4] | 37.3 [36.7-37.9] |
| Female (%, [95% CI]) | 50.5 [50.1-50.8] | 60.9 [60.4-61.4] | 50.1 [49.4-50.7] | 50.3 [48.9-51.8] | 59.5 [58.7-60.2] | 49.5 [48.0-51.0] | 57.4 [55.7-59.0] | 52.0 [50.2-53.7] |
| Pain (/10, median [IQR]) | 4 [0-7] | 6 [2-8] | 0 [0-5] | 5 [2-8] | 4 [0-7] | 4 [0-6] | 7 [4-8] | 4 [0-7] |
| Acuity (/5, median [IQR]) | 2 [2-3] | 3 [2-3] | 3 [2-3] | 4 [4-4] | 3 [3-3] | 4 [4-4] | 4 [4-4] | 4 [4-4] |
| Length of stay (mins, mean [95% CI]) | 458 [456-461] | 414 [411-417] | 297 [297-300] | 214 [209-219] | 261 [259-264] | 157 [154-160] | 213 [207-219] | 151 [148-155] |
| Discharged home (%, [95% CI]) | 37.0 [36.7-37.3] | 74.9 [74.5-75.3] | 91.4 [91.0-91.8] | 95.3 [94.6-95.8] | 86.6 [86.1-87.1] | 95.4 [94.8-96.0] | 96.7 [96.1-97.2] | 87.0 [85.8-88.1] |

[1] 95% confidence intervals (CI) are cluster-robust at the patient level (intercept-only GEE with robust sandwich standard errors); patients may contribute more than one visit

[2] Only female/male are reported in the data

[3] Emergency Severity Index, 1 = most severe

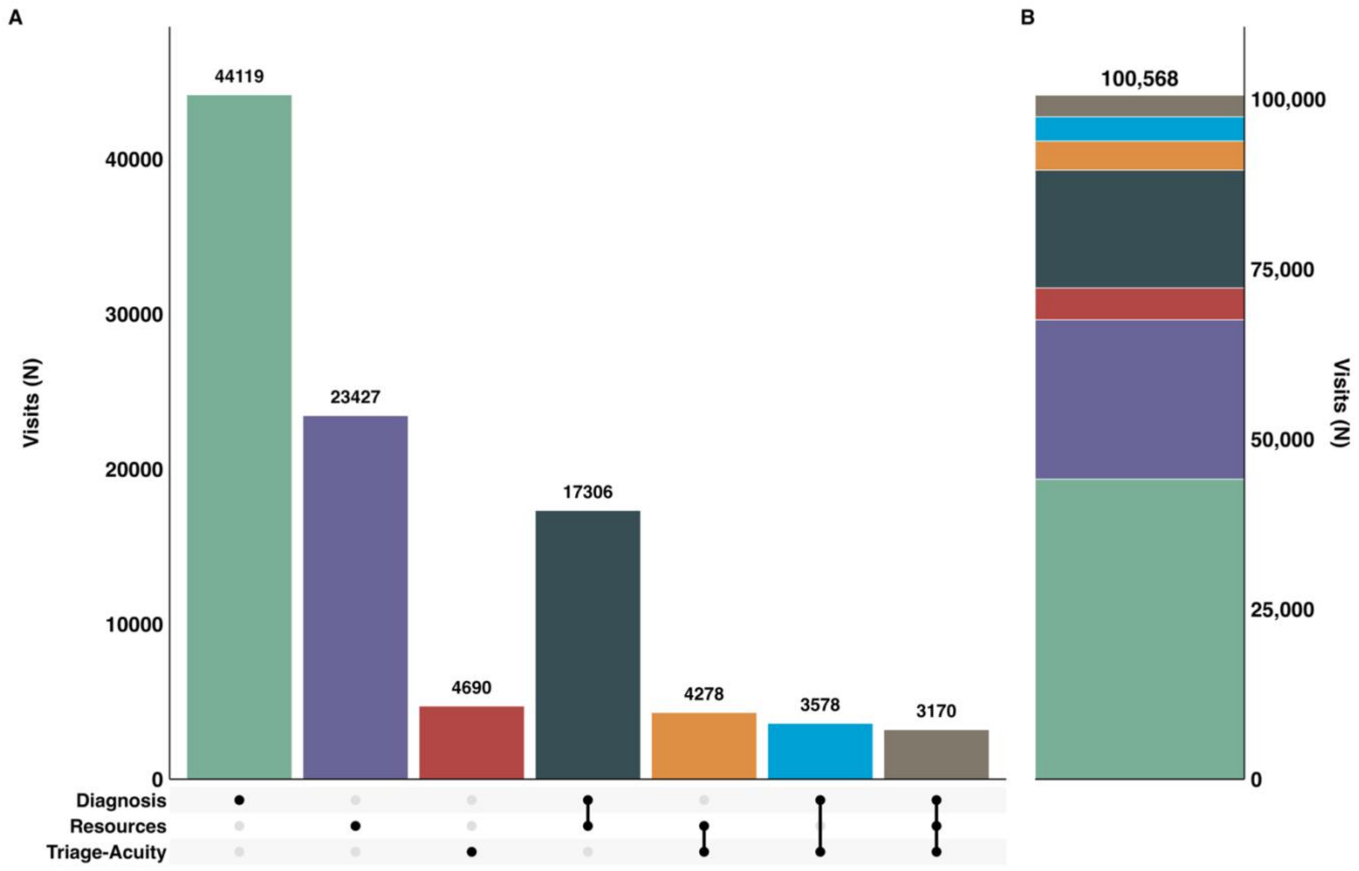


**Figure S3.** The counts and intersections between the potentially avoidable ED visits identified by each classification algorithm in the sensitivity analysis sample (1 visit per patient).

**Table S8.** Three most common chief complaints, overall and per algorithm combination (one visit per patient)

| Rank | Overall | Diagnosis | Resource | Triage Acuity | Diagnosis+ Resources | Resources+ Triage Acuity | Diagnosis+ Triage Acuity | All |
|---|---|---|---|---|---|---|---|---|
| 1st | Abdomen pain (6.0%) | Abdomen pain (10.2%) | Alcohol intoxication (9.7%) | Finger laceration (10.9%) | Abdomen pain (9.3%) | Finger laceration (12.4%) | Lower back pain (8.8%) | Rash (10.7%) |
| 2nd | Chest pain (4.4%) | Nausea vomiting (9.8%) | Syncope (6.2%) | Ankle pain (9.1%) | Nausea vomiting (4.4%) | Ankle pain (9.9%) | Back pain (7.3%) | Ear pain (7.2%) |
| 3rd | Nausea vomiting (4.2%) | Headache (5.5%) | Chest pain (5.4%) | Thumb laceration (7.2%) | Leg pain (4.3%) | Thumb laceration (8.3%) | Sore throat (6.0%) | Eye pain (7.1%) |

**Table S9.** Three most common discharge diagnosis categories, overall and per algorithm combination (one visit per patient)

| Rank | Overall | Diagnosis | Resource | Triage Acuity | Diagnosis+ Resources | Resources+ Triage Acuity | Diagnosis+ Triage Acuity | All |
|---|---|---|---|---|---|---|---|---|
| 1st | Ill-defined conditions (23.1%) | Ill-defined conditions (44.6%) | Injury and poisoning (31.5%) | Injury and poisoning (82.8%) | Ill-defined conditions (51.6%) | Injury and poisoning (80.3%) | Diseases of the musculoskeletal system (43.0%) | Diseases of the musculoskeletal system (29.6%) |
| 2nd | Injury and poisoning (21.2%) | Diseases of the musculoskeletal system (15.3%) | Ill-defined conditions (27.5%) | Diseases of the musculoskeletal system (4.0%) | Diseases of the musculoskeletal system (16.8%) | Mental disorders (3.8%) | Diseases of the skin and subcutaneous tissue (10.5%) | Ill-defined conditions (16.8%) |
| 3rd | Diseases of the circulatory system (7.9%) | Diseases of the skin and subcutaneous tissue (7.7%) | Mental disorders (17.6%) | Factors influencing health status/contact with health services (2.3%) | Diseases of the genitourinary system (5.3%) | Factors influencing health status/contact with health services (3.2%) | Ill-defined conditions (10.3%) | Diseases of the skin and subcutaneous tissue (10.4%) |

The Cohen’s Kappa values for agreement Diagnosis x Resources = 0.039 [0.036–0.042], Diagnosis x Triage-Acuity = 0.099 [0.095-0.103], Resources x Triage-Acuity = 0.131 [0.126-0.135]

There were few meaningful differences between the analysis performed on the analysis sample and the sensitivity sample.

### Sensitivity analysis 3 — Triage-Acuity defined as ESI 5 only

**Table S10.** Characteristics of the sensitivity analysis sample (Triage-Acuity ESI=5)[1]

| | |
|---|---|
| **Age (Years, mean)** | 53 [53, 53] |
| **(Years, median)** | 53 (18-103) |
| **Sex (%)** | |
| Female | 54% [54%, 54%] |
| Male | 46% [46%, 46%] |
| **Self-reported Race (%)[2]** | |
| White | 58% [58%, 58%] |
| Black | 22% [22%, 22%] |
| Hispanic/Latino | 8% [8.1%, 8.2%] |
| Other | 7% [7.3%, 7.4%] |
| Asian | 4% [4.3%, 4.4%] |

[1]Confidence interval (CI) for categorical variables is computed using Agresti-Coull interval.

[2]For clarity, self-reported races have been collapsed.

Abbreviation: CI = Confidence Interval

**Table S11.** Potentially avoidable visit categories (Triage-Acuity ESI=5)

| Measure | Not identified by any algorithm | Potentially Avoidable ED Visits (by Algorithm) | | | | | | |
|---|---|---|---|---|---|---|---|---|
| | | Diagnosis | Resource | Triage Acuity | Diagnosis+ Resources | Resources+ Triage Acuity | Diagnosis+ Triage Acuity | ALL |
| Number of visits (N) | 216,233 | 108,893 | 50,207 | 153 | 41,292 | 260 | 157 | 528 |
| Proportion (%, [95% CI])[1] | 51.8 [51.6-51.9] | 26.1 [25.9-26.2] | 12.0 [11.9-12.1] | <0.1 [0.03-0.04] | 9.9 [9.8-10.0] | <0.1 [0.03-0.04] | 0.1 [0.1-0.1] | 0.1 [0.1-0.1] |
| Age (years, mean [95% CI])[1] | 56.6 [56.5-56.7] | 50.6 [50.5-50.7] | 46.2 [46.0-46.4] | 42.6 [39.7-45.4] | 47.3 [47.1-47.5] | 39.8 [37.6-42.0] | 39.7 [37.3-42.2] | 38.9 [37.5-40.4] |
| Female (%, [95% CI])[1,2] | 51.2 [51.0-51.4] | 62.1 [61.8-62.4] | 47.9 [47.5-48.3] | 47.7 [40.0-55.6] | 58.0 [57.5-58.5] | 49.6 [43.6-55.7] | 51.6 [43.8-59.3] | 45.5 [41.3-49.7] |
| Pain (/10, median [IQR]) | 4 [0-8] | 7 [2-9] | 1 [0-6] | 1 [0-6] | 4 [0-7] | 0 [0-3] | 4 [0-8] | 0 [0-3] |
| Acuity (/5, median [IQR])[3] | 2 [2-3] | 3 [2-3] | 3 [2-3] | 5 [5-5] | 3 [3-3] | 5 [5-5] | 5 [5-5] | 5 [5-5] |
| Length of stay (mins, mean [95% CI])[1] | 491 [489-493] | 433 [431-435] | 301 [299-303] | 263 [209-317] | 252 [251-254] | 128 [115-141] | 181 [150-212] | 104 [97-111] |
| Discharged home (%, [95% CI])[1] | 37.0 [36.8-37.2] | 72.3 [72.0-72.5] | 90.1 [89.9-90.4] | 90.8 [85.1-94.6] | 85.0 [84.6-85.3] | 85.4 [80.5-89.2] | 94.3 [89.3-97.1] | 82.8 [79.3-85.8] |
| Admitted to hospital (%, [95% CI])[1] | 58.4 [58.2-58.6] | 25.0 [24.7-25.2] | NA[4] | 5.9 [3.0-10.9] | NA[4] | NA[4] | <0.1 [0.00-2.87] | NA[4] |

[1]95% confidence intervals (CI): Agresti-Coull for proportions; t-distribution for means

[2]Only female/male are reported in the data

[3]Emergency Severity Index, 1 = most severe

[4]NA; Not applicable. The Resource algorithm excludes visits that were admitted by design

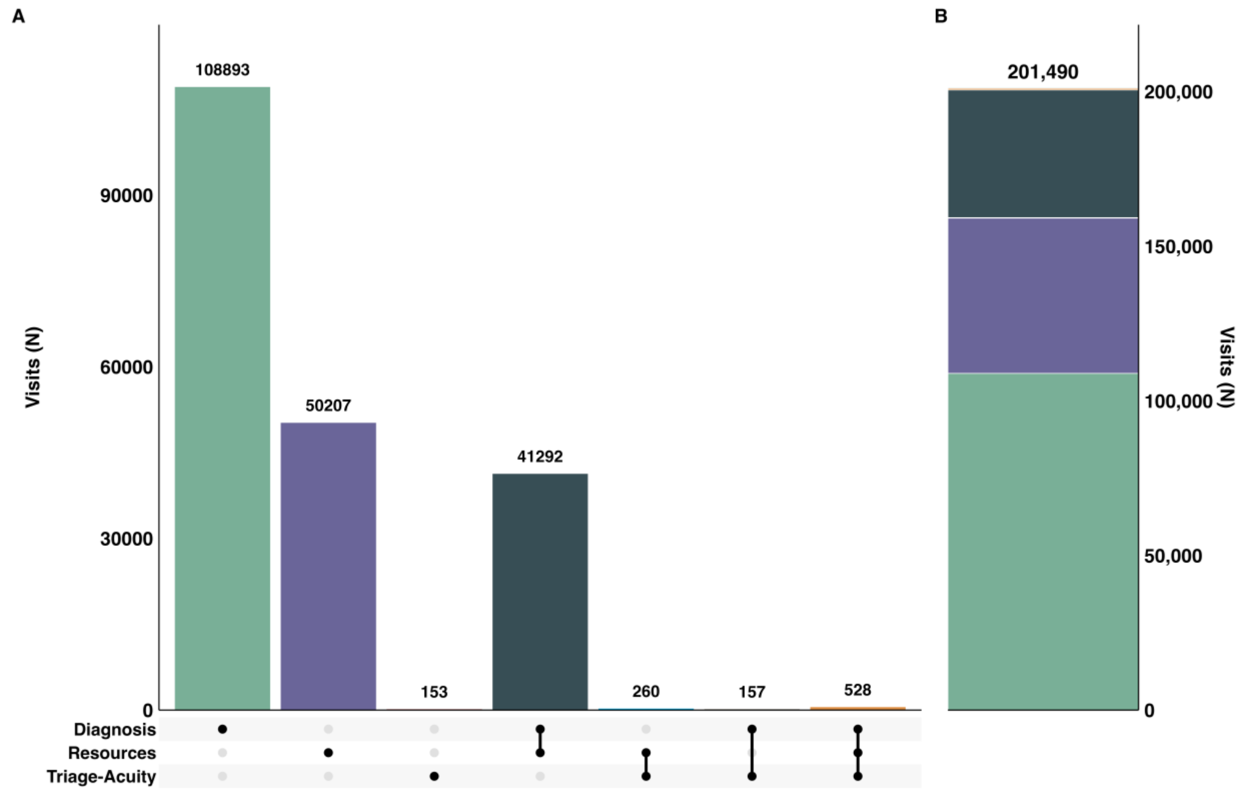


**Figure S4.** The counts and intersections between the potentially avoidable ED visits identified by each classification algorithm, with Triage-Acuity defined as ESI 5 only.

**Table S12.** Three most common chief complaints, overall and per algorithm combination (Triage-Acuity ESI=5)

| Rank | Overall | Diagnosis | Resource | Triage Acuity | Diagnosis+ Resources | Resources+ Triage Acuity | Diagnosis+ Triage Acuity | All |
|---|---|---|---|---|---|---|---|---|
| 1st | Abdomen pain (5.5%) | Nausea vomiting (9.7%) | Alcohol intoxication (10.0%) | Unknown (15.0%) | Abdomen pain (7.6%) | Unknown (18.1%) | Unknown (17.2%) | Laceration (27.3%) |
| 2nd | Chest pain (5.1%) | Abdomen pain (9.6%) | Chest pain (5.1%) | Subarachnoid hemorrhage transfer (11.1%) | Nausea vomiting (4.2%) | Laceration (11.2%) | Dental pain (12.7%) | Unknown (21.6%) |
| 3rd | Dyspnea (4.4%) | Chest pain (5.2%) | Syncope (4.1%) | Finger laceration (9.8%) | Leg pain (4.2%) | Subarachnoid hemorrhage transfer (7.7%) | Foot pain (7.6%) | Wound evaluation (7.0%) |

**Table S13.** Three most common discharge diagnosis categories, overall and per algorithm combination (Triage-Acuity ESI=5)

| Rank | Overall | Diagnosis | Resource | Triage Acuity | Diagnosis+ Resources | Resources+ Triage Acuity | Diagnosis+ Triage Acuity | All |
|---|---|---|---|---|---|---|---|---|
| 1st | Ill-defined conditions (24.0%) | Ill-defined conditions (42.2%) | Injury and poisoning (33.5%) | Injury and poisoning (56.2%) | Ill-defined conditions (46.5%) | Injury and poisoning (52.7%) | Factors influencing health status/contact with health services (29.9%) | Factors influencing health status/contact with health services (50.2%) |
| 2nd | Injury and poisoning (16.7%) | Diseases of the musculoskeletal system (17.6%) | Ill-defined conditions (24.3%) | Factors influencing health status/contact with health services (15.0%) | Diseases of the musculoskeletal system (19.5%) | Factors influencing health status/contact with health services (17.3%) | Diseases of the musculoskeletal system (22.9%) | Diseases of the musculoskeletal system (11.6%) |
| 3rd | Diseases of the digestive system (8.2%) | Diseases of the skin and subcutaneous tissue (7.5%) | Mental disorders (18.3%) | Mental disorders (7.2%) | Diseases of the skin and subcutaneous tissue (5.0%) | Mental disorders (8.8%) | Diseases of the digestive system (12.1%) | Diseases of the skin and subcutaneous tissue (7.8%) |
